# Widespread aberrant alternative splicing despite molecular remission in chronic myeloid leukemia patients

**DOI:** 10.1101/2020.07.31.20165597

**Authors:** Ulf Schmitz, Jaynish S. Shah, Bijay P. Dhungel, Geoffray Monteuuis, Phuc-Loi Luu, Veronika Petrova, Cynthia Metierre, Shalima S. Nair, Charles G. Bailey, Verity A. Saunders, Ali G. Turhan, Deborah L. White, Susan Branford, Susan Clark, Timothy P. Hughes, Justin J-L. Wong, John E.J. Rasko

**Author notes:** **Correspondence:** John E.J. Rasko, Locked Bag No. 6, Newtown 2042, NSW Australia.

## Abstract

**Background:** Vast transcriptomics and epigenomics changes are characteristic of human cancers including leukemia. At remission, we assume that these changes normalise so that omics-profiles resemble those of healthy individuals. However, an in-depth transcriptomic and epigenomic analysis of cancer remission has not been undertaken. A striking exemplar of targeted remission induction occurs in chronic myeloid leukemia (CML) following tyrosine kinase inhibitor (TKI) therapy.

**Methods:** Using RNA sequencing and whole-genome bisulfite sequencing, we profiled samples from chronic-phase CML patients at diagnosis and remission, and compared these to healthy donors.

**Results:** Remarkably, our analyses revealed that abnormal splicing distinguishes remission samples from normal controls. This phenomenon is independent of the TKI drug used and in striking contrast to the normalisation of gene expression and DNA methylation patterns. Most remarkable are the high intron retention (IR) levels that even exceed those observed in the diagnosis samples. Increased IR affects cell cycle regulators at diagnosis and splicing regulators at remission. We show that aberrant splicing in CML is associated with reduced expression of specific splicing factors, histone modifications and reduced DNA methylation.

**Conclusions:** Our results provide novel insights into the changing transcriptomic and epigenomic landscapes of CML patients during remission. The conceptually unanticipated observation of widespread aberrant alternative splicing after remission induction warrants further exploration. These results have broad implications for studying CML relapse and treating minimal residual disease.

## Background

Characterisation of *BCR-ABL1* and its constitutive tyrosine kinase activity has facilitated understanding of the pathogenesis and therapy of CML^1^. Tyrosine kinase inhibitor (TKI) treatment results in positive responses in 60-90% of patients with chronic phase CML^2,3^. However, molecular mechanisms that facilitate CML maintenance and relapse remain elusive^4^. While the molecular causes of CML are well understood, measures of therapy success based on *BCR-ABL1* expression and white blood cell count are not reliable predictors of relapse.

*BCR-ABL1* overexpression perturbs the hematopoietic transcriptome^4,5^. CML progression, maintenance, and therapy may be affected by aberrant expression and alternative splicing of cancer-causing genes^6,7^. Alternative splicing is vital for cellular homeostasis and dysregulated intron retention (IR) has been associated with numerous human cancers including leukemia^8^. However, the role of alternative splicing in CML remains unknown.

In addition to risk-associated genomic variants which predict poor outcome^9^, there is evidence that epigenetic regulation affects CML pathology and therapy success^10^. Epigenetic regulation via DNA methylation and nucleosome occupancy play key roles in constitutive and alternative mRNA splicing regulation^11-13^. Epigenetic regulation via DNA methylation is known to regulate constitutive and alternative mRNA splicing. To date, no systematic transcriptomics and epigenomics analyses in CML remission samples have been conducted, neither have the effects of different therapeutic regimens on gene expression and alternative splicing been assessed. CML presents an excellent model to study epigenetically mediated transcriptomic alterations in myeloid cells that cause malignant transformation and the response to TKI treatment. New insights into this multilayered regulatory network could provide important clues for targeted therapies used in other malignancies as well.

Here we report novel transcriptional and epigenetic patterns in a multi-omics analysis of CML patient samples before and after effective front-line TKI treatment. Using this model, we found abnormal splicing in chronic-phase CML patients, which we confirmed in a larger independent cohort. Surprisingly, we also observed aberrant splicing patterns in complete molecular remission. This is in contrast to the normalisation of gene expression and DNA methylation patterns driven by the reconfiguration of blood cell composition. More specifically, we observed a marked increase in IR and differential exon usage in CML diagnosis and remission compared to normal samples, as well as evidence that epigenetic factors modulate splicing in CML.

## Methods

### Patient samples

We retrieved nine diagnostic specimens (total leukocytes from peripheral blood collected in EDTA) from patients enrolled in the RESIST (1x imatinib; ACTRN12610000055000), ENESTxtnd (4x nilotinib, NCT01254188) and PINNACLE (4x nilotinib + pegylated interferon, ACTRN12612000851864) trials, with matched remission specimens (major or complete molecular response at 12 or 24 months). In addition, eight CML patient samples (7 diagnostic and 1 matched remission) from the French Persistem study were obtained. For the validation of IR events and fusion transcripts we retrieved further matched diagnosis/remission samples from the TIDELII trial (20x imatinib, ACTRN12607000325404). RNA was isolated from mononuclear cells (from peripheral blood collected in LiHep) which had been cryopreserved and subsequently thawed into TRIzol. All studies were approved by the institutional review boards (HREC protocol No: 131015, 070718c, 081211, and 101010 - Royal Adelaide Hospital) and are in accordance with the Declaration of Helsinki. Patients provided written informed consent.

### RNA isolation and mRNA-seq

Total RNA was isolated from diagnosis, remission, and control samples (peripheral blood mononuclear cells - PBMCs) using Trizol according to the manufacturer’s protocol. For mRNA sequencing, poly-A enriched mRNA libraries were prepared from 1 μg of total RNA using the TruSeq Stranded RNA sample prep kit (Illumina), prior to paired-end sequencing using the HiSeq 2500 platform. RNA sequencing was performed in triplicates for each sample. The raw sequencing data (fastq files) have been deposited at GEO under accession GSE144119.

### mRNA sequencing data analysis

Paired-end RNA-sequencing reads (125 nt) were trimmed and mapped to the human reference genome hg38 using STAR^14^. Quality control of raw and mapped sequencing reads was performed using FASTQC (github.com/s-andrews/FastQC), RSeQC^15^, and multiqc^16^. Putative confounders and batch effects were excluded using principal components analyses (Supplementary Figure 1). STAR-FUSION^17^ was used for the identification of fusion genes and Fusion Inspector (FusionInspector.github.io) for *in silico* validation of the predicted gene fusions. Gene expression levels specified as transcripts per million (TPM) were determined using Salmon 0.14.1^18^.

General statistics on alternative splicing events were determined using the rMATS software^19^. We used our IRFinder algorithm for the detection of IR events in introns extracted from an Ensembl gtf file (regions between two adjacent exons)^20^. IRFinder estimates the abundance of IR by computing the ratio between gene transcripts retaining an intron and the sum of all transcripts of the respective gene (more information is provided in the Supplementary Materials).

Differentially used exons were determined using the R Bioconductor package DEXSeq^21^. Differential exon usage (DEU) = changes in the relative usage of exons:

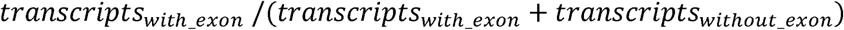

### IR validation

Extracted RNA (2 µg) was treated with DNAse Turbo (Invitrogen) and reverse transcribed into cDNA using oligo(dT) priming and Superscript III (ThermoFisher) according to the protocol supplied by the manufacturer. For each sample, a corresponding control without reverse transcriptase was used to assess DNA contamination. For qPCR, cDNA templates were amplified, and the Ct values were quantified with SYBR Green Master Mix (ThermoFisher). Beta-2 microglobulin (*B2M*) was used as the normalization control for cDNA input. Experiments were performed using the CFX96 Real-Time PCR System (BIORAD). The list of primers is provided in Supplementary Table 1.

### Whole genome bisulfite sequencing (WGBS)

WGBS libraries were prepared following Illumina’s “Whole-Genome Bisulfite sequencing for Methylation Analysis” protocol. Briefly, 1 μg of genomic DNA was spiked with 0.5% unmethylated lambda DNA and sonicated to generate fragments of size between 150 to 300 bp. Library preparation was performed using Illumina’s Paired-end DNA Sample Prep Kit (discontinued, Illumina, CA, USA) according to the manufacturer’s protocol. The size-selected libraries were then subjected to bisulfite conversion as previously described^22^. Adaptor-ligated bisulfite treated DNA was enriched by 10 cycles of PCR amplification using the PfuTurbo Cx Hotstart DNA Polymerase (Stratagene). Qualitative and quantitative checks of the libraries were performed using Agilent’s High sensitivity DNA kit (Agilent) and KAPA Library quantification kit (KAPA Biosystems). Three lanes of paired-end 100 bp sequencing was performed for each of the library on the Illumina HiSeq2500 platform using the TruSeq v3 cluster kits and SBS kits to achieve coverage ranging between 25-30x.

### WGBS data analysis

Reads were processed and aligned to the human (hg38) reference genome using Meth10X (github.com/luuloi/Meth10X)^23^. In short, the Meth10X pipeline takes raw reads in fastq format and trims the adaptors, which are then aligned to the human reference genome using bwa-meth (github.com/brentp/bwa-meth). The generated bam files were marked with duplication and merged if necessary. Estimation of the duplication rate, coverage bias (genomic features) and methylation bias in reads was carried out to provide quality control. All the metrics such as percentage of unmapped/mapped read metrics, mapping quality distribution, GC content distribution, insert size distribution and coverage distribution were generated by Qualimap 2 for further evaluation of the alignment. Finally, a count table and bigwig files of methylated and coverage at each CpG site in the genome was constructed for all samples. Differentially methylated regions (DMRs) were identified using MethPipe^24^ with count table of all samples as input. The analysis of differential methylation and IR is described in the Supplementary Materials.

### ChIP-seq data analysis

ChIP-seq data of histone modifications in K562 cells was retrieved from ENCODE (encodeproject.org) and further processed as described in the Supplementary Materials.

### Statistical analyses

All statistical analyses were performed in R v.3.6.2. The Wald test implemented in the R package DESeq2^25^ was used for the identification of statistically significant differential alternative splicing events, and differential gene expression analysis. P-values were adjusted for the false discovery rate using the Benjamini-Hochberg procedure. Functional enrichment analysis of differentially expressed genes and genes with differentially retained introns was performed using DAVID 6.7^26^ with the human genome and the set of expressed genes, respectively, used as a background.

## Results

The goal of our work was to evaluate transcriptomic and epigenetic changes in CML patients after major or complete molecular response. We retrieved sixteen peripheral blood samples of Philadelphia-positive (Ph+) CML patients. For 10 of these patients, we also retrieved matched samples after remission. In addition, we retrieved peripheral blood from six healthy donors (Table 1). We subjected all samples to RNA sequencing and all matched diagnosis/remission samples as well as control samples to WGBS.

**Table 1.**
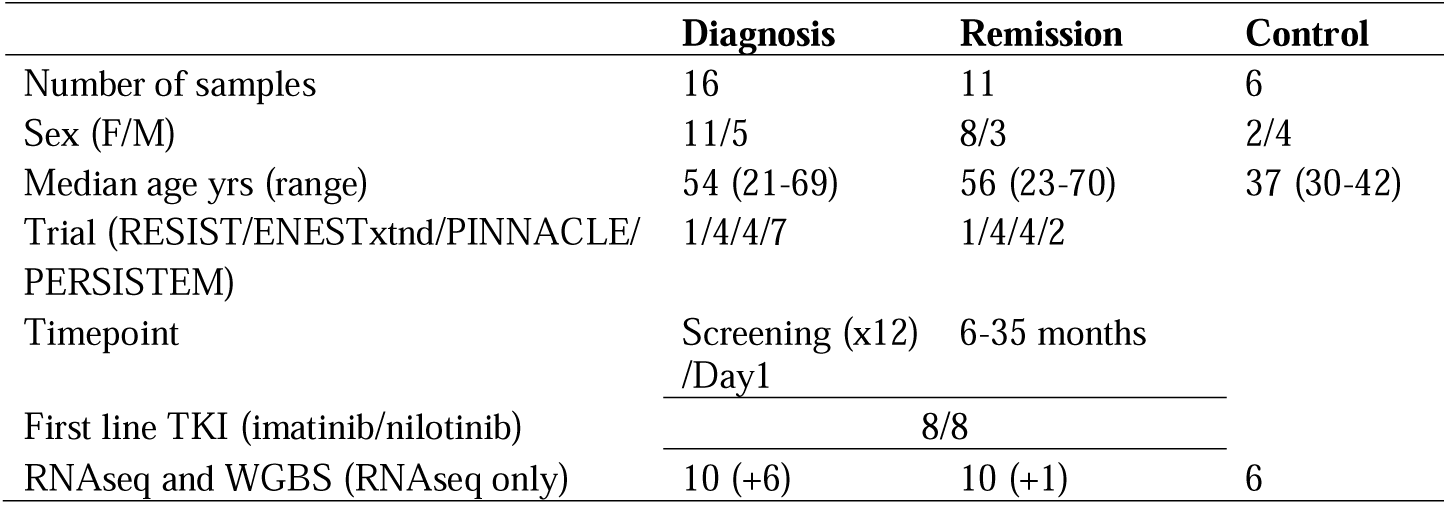
Patient and healthy donor samples used in RNAseq and WGBS experiments. Sequencing was performed on PBMCs.

### Heterogeneous CML transcriptomes converge at remission

RNA sequencing reads mapping to the *BCR-ABL1* locus were found in diagnosis samples only (Supplementary Figure 2A), confirming the results of clinical qRT-PCR-based *BCR-ABL1* quantification (Supplementary Data 1). Apart from the characteristic *BCR-ABL1* fusion we identified other recurring intra- and inter-chromosomal fusions (Supplementary Figure 2A&B; Supplementary Data 2) that occur in diagnosis, remission or control samples. A novel fusion transcript between the myeloid cell-specific transmembrane glycoprotein *CLEC12A* (12p13.31) and the microRNA *miR-223* host gene *MIR223HG* (Xq12) was expressed at higher levels (*FC* = 7.16) at diagnosis compared to remission (Supplementary Figure 2C&D). For most patients we observed a global reduction of gene expression in remission compared to diagnosis (Supplementary Figure 3A). This can be attributed to the reconfigured blood cell composition, which resembles lymphoid cell-enriched normal controls (Figure 1A&B). A significant downregulation was observed in genes encoding kinases other than *BCR-ABL1* and kinase-like proteins such as *CENPE, CDK1, AURKB, MELK*, and *BUB1B* (Supplementary Figure 4). Reduced expression at remission, similar to levels observed in control samples, also affected cyclins such as *CCNA1, CCNB2* and other cell cycle regulators as well as genes related to DNA replication and repair (Figure 1A & Supplementary Figure 5A). In this context, no consistent differences were noted between patients treated with imatinib or nilotinib (Supplementary Figure 1).

**Figure 1.**
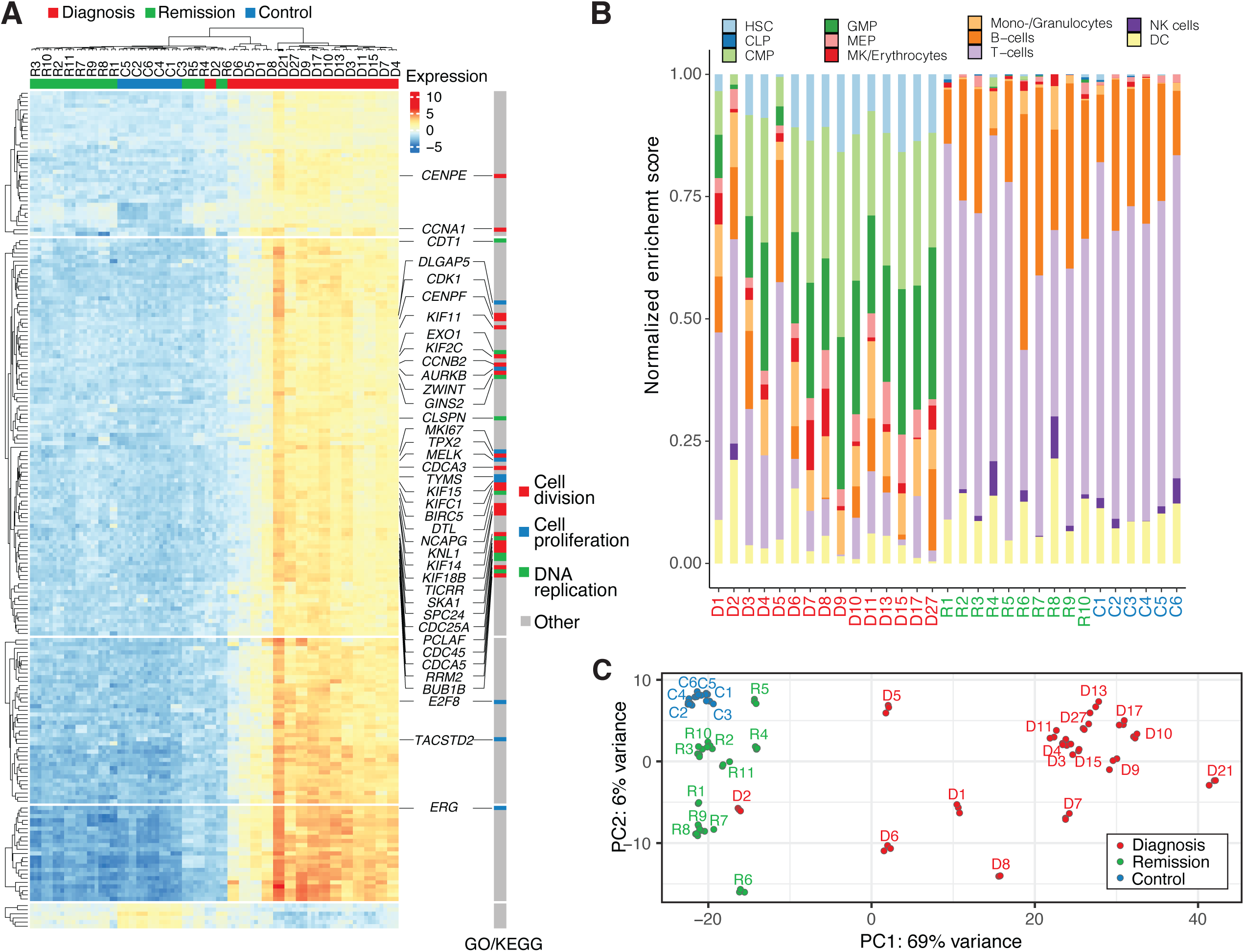
Molecular remission is reflected in transcriptomic profiles. **(A)** The Top 200 differentially expressed genes with lowest adjusted *p*-value in likelihood ratio test. **(B)** Predicted cell type enrichment in peripheral blood samples. The stacked bar graph shows normalized cell type enrichment scores for patient and control transcriptomes based on bulk RNA sequencing data. Predictions were made with the *xCell* R package (github.com/dviraran/xCell)^47^. HSC – hematopoietic stem cell, CLP – common lymphoid progenitor, CMP – common myeloid progenitor, GMP - granulocyte macrophage progenitor, MEP - megakaryocytic erythroid progenitor, NK – natural killer, DC – dendritic cell. **(C)** Principal component analysis separates diagnosis samples from the common features of remission samples and healthy controls.

A set of genes (n=81), that are functionally associated with oxygen and bicarbonate transport (GO:0015671; GO:0015701) as well as the hemoglobin’s chaperone pathway (Biocarta), was significantly differentially expressed in all three comparisons (diagnosis vs control, remission vs control, and diagnosis vs remission; Supplementary Figure 5B). Overall, we observed that patients diagnosed with CML have highly heterogeneous transcriptomes. In contrast, remission transcriptomes are more alike (Figure 1C) and exhibit fewer differences in comparison to healthy control samples (Supplementary Figure 3B).

### DNA methylation profiles return to normal at remission

To characterise putative epigenetic causes for differential gene expression in CML patients prior to and after successful TKI treatment, all matched diagnosis/remission samples as well as all control samples were subjected to WGBS. The analysis revealed almost 25,000 differentially methylated regions (DMRs) in the genomes of diagnosis and remission samples (Figure 2A, left). Vast DNA methylome differences were found in every matched patient sample (Supplementary Figure 6) and across all chromosomes (Figure 2B). In contrast, only 710 DMRs were observed between remission and control samples (Figure 2A, right). Indeed, multidimensional scaling showed that DNA methylation profiles of remission samples are closer to those from healthy donors rather than to matched diagnosis samples (Supplementary Figure 7). DMRs between diagnosis and remission samples contain, on average, a larger number of differentially methylated CpG sites (Figure 2C). This is consistent with previous reports that suggest an increase in CpG site methylation during CML progression that affects tumour-suppressor genes and regulators of cell proliferation^10^. DMRs are associated with processes of the innate immune response, apoptosis, cell differentiation, and cell migration (Figure 2D).

**Figure 2.**
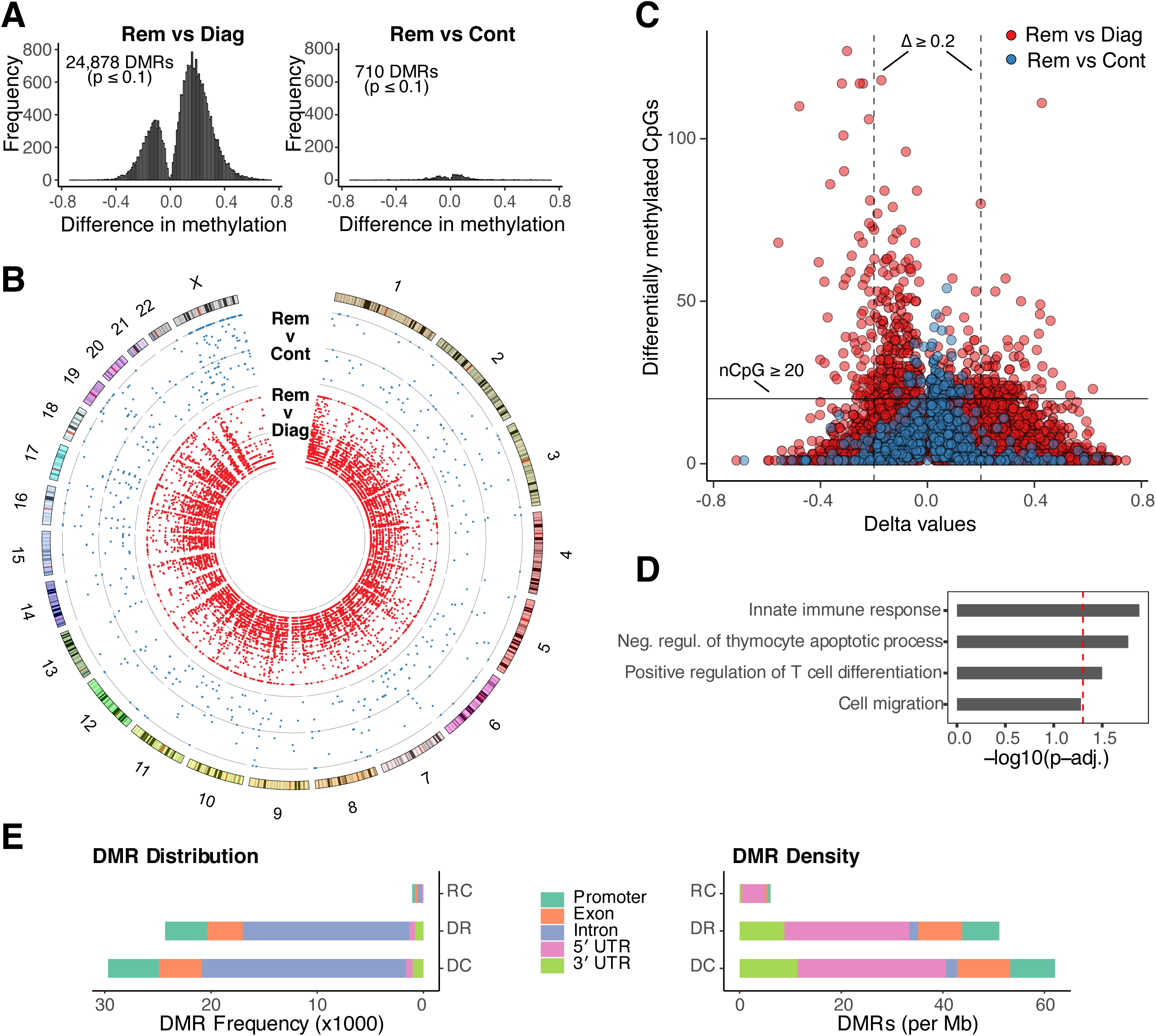
DNA methylation in CML. **(A)** Frequency of differentially methylated regions (DMRs) based on delta values between pooled diagnosis and remission samples (left) as well as remission and control samples (right). **(B)** Circos plot illustrating genome-wide differences in the frequency of DMRs (central ring with blue scatter plot: diagnosis vs remission; inner ring with red scatterplot: remission vs control). This plot was generated using the Circos software^48^. **(C)** Scatterplot illustrating the number of significantly differentially methylated CpG sites per DMR and associated delta values (red points, diagnosis vs remission; blue points, remission vs control). **(D)** Enriched GO terms associated with DMRs (remission vs diagnosis) in gene promoter regions (red dashed line marks *p* = 0. 05). **(E)** Distribution and density of DMRs across gene features. RC – remission vs control; DR – diagnosis vs remission; DC – diagnosis vs control.

Our results show that DNA methylation returns to normal levels after successful TKI treatment. Most DMRs can be found in intronic regions, however, the highest density of DMRs was observed in 5′ untranslated regions (Figure 2E), suggesting a key role for differential DNA methylation in orchestrating CML gene expression regulation.

### Aberrant alternative splicing distinguishes remission samples from healthy controls

To assess whether major or complete molecular remission also affects gene isoform expression, we conducted a systematic analysis of the five major forms of alternative splicing (Supplementary Figure 8A). In a pairwise comparison of alternative splicing event frequencies between matched diagnosis/remission samples, no consistent trend towards an increase or decrease in either of the two conditions was found (Supplementary Figure 8B). This suggests, that in contrast to gene expression profiles, splicing patterns remain atypical after major or complete molecular remission.

Alternative splicing analysis of the Cancer Genome Atlas (TCGA) cohort by Dvinge et al. (2015) revealed that the frequency of IR events is consistently higher in acute myeloid leukemia compared to normal controls^8^. Although mean IR frequencies are also higher in CML samples (diagnosis, 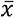 = 3 382; remission, 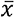 = 3 484) compared to healthy donors (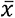 = 2 822), high inter-donor variability led to differences in IR frequencies that did not reach significance (Supplementary Figure 8C). However, when we subsampled the RNA sequencing data to facilitate a uniform sequencing depth for comparison, we observed a clear separation with significantly increased IR frequencies at diagnosis and remission compared to controls (Supplementary Figure 9). Moreover, we found that the IR ratios are consistently higher in diagnosis and remission samples compared to healthy controls (Figure 3A). Differences between diagnosis and remission samples were less pronounced (Figure 3A; Supplementary Figure 10). While IR profiles are unsuitable for patient stratification (Supplementary Figure 11), differentially retained introns between diagnosis and remission samples are, similarly to differentially expressed genes, associated with cell cycle regulators as well as genes related to DNA replication and repair (Supplementary Figure 12).

**Figure 3.**
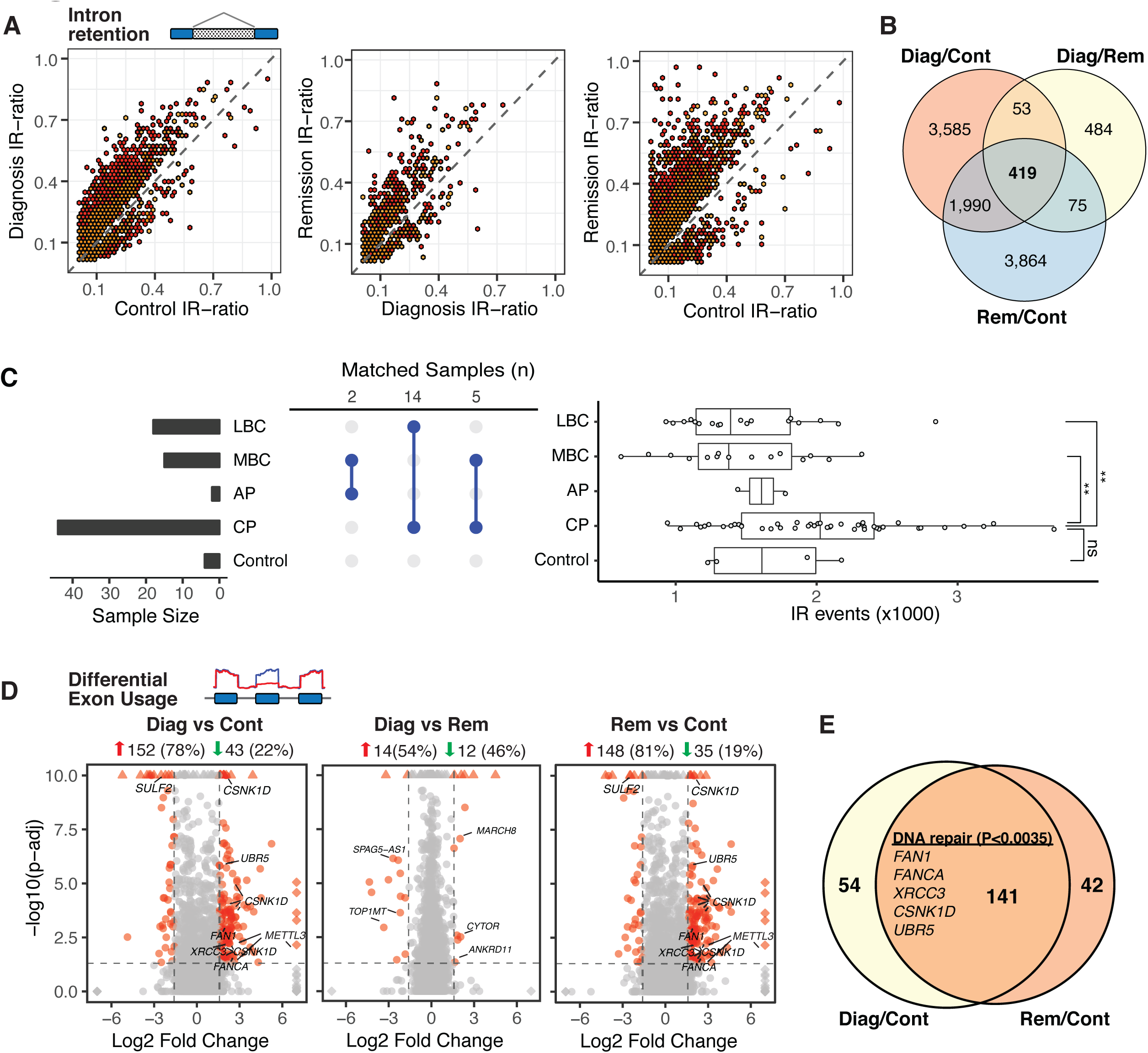
Global alternative splicing patterns are similar despite major molecular remission. **(A)** Scatterplots illustrating differential IR events between pooled diagnosis/remission/control samples binned into hexagons to avoid overplotting (yellow: *p_adj_* <= 0.05; red: *p_adj_* <= 0.01). **(B)** Venn diagram showing intersections of mutually differentially retained introns. **(C)** Dynamics of IR across CML phases. CML patient data from Branford et al.^9^ was analysed for the occurrence of IR events. Left: Sample sizes for Control, CP: Chronic Phase, AP: Accelerated Phase, MBC: Myeloid Blast Crisis, and LBC: Lymphoid Blast Crisis samples. Middle: Sample numbers matched from different phases. Right: Boxplot of IR frequencies in samples from different CML phases. Mann–Whitney U test; ** (p < 0.01), ns (p > 0.05). **(D)** Differential exon usage in pooled diagnosis vs control (left), diagnosis vs remission (middle), and remission vs control samples. **(E)** Intersection of differentially used exons in diagnosis and remission samples compared to controls (the top-enriched GO term and associated genes are listed in the middle).

However, in remission IR primarily affects genes involved in splicing processes (Supplementary Figure 12) suggesting the aberrant splicing persists in remission due to auto-regulatory processes. Overall, IR profiles are highly dynamic in CML, which becomes apparent when considering the large overlap of differentially retained introns (n = 419) in all three comparisons (Figure 3B). We confirmed these results with an independent dataset of 59 CML patients from Branford et al.^9^, which includes samples from various stages of disease progression, and from 4 healthy controls (Figure 3C). The number of IR events in this cohort was highly variable. While there are on average more IR events in chronic phase compared to healthy controls, the number drops significantly at blast crisis (Figure 3C, right).

Examining differential exon usage in our samples we observed a similar trend although not as marked as observed in differential IR, i.e. both diagnosis and remission samples exhibit a larger number of differentially used exons (DUEs) compared to normal controls. Most of these DUEs are upregulated (diagnosis, 78%; remission, 81%; Figure 3D). A large fraction of the DUEs at diagnosis (72%) remain differentially expressed at remission as well. Some genes of these mutual DUEs are associated with DNA repair mechanisms (top-enriched GO term; Figure 3E). Only 26 exons in total are differentially used in diagnosis vs remission samples (Figure 3D).

### Modulation of IR levels supports lineage-specific gene expression in CML

IR is accepted as a mechanism of post-transcriptional gene regulation triggering nonsense-mediated decay due to the presence of premature termination codons within the retained introns^27,28^. We analysed the relationship between IR and gene expression changes but overall the two variables did not significantly correlate (Figure 4A).

**Figure 4.**
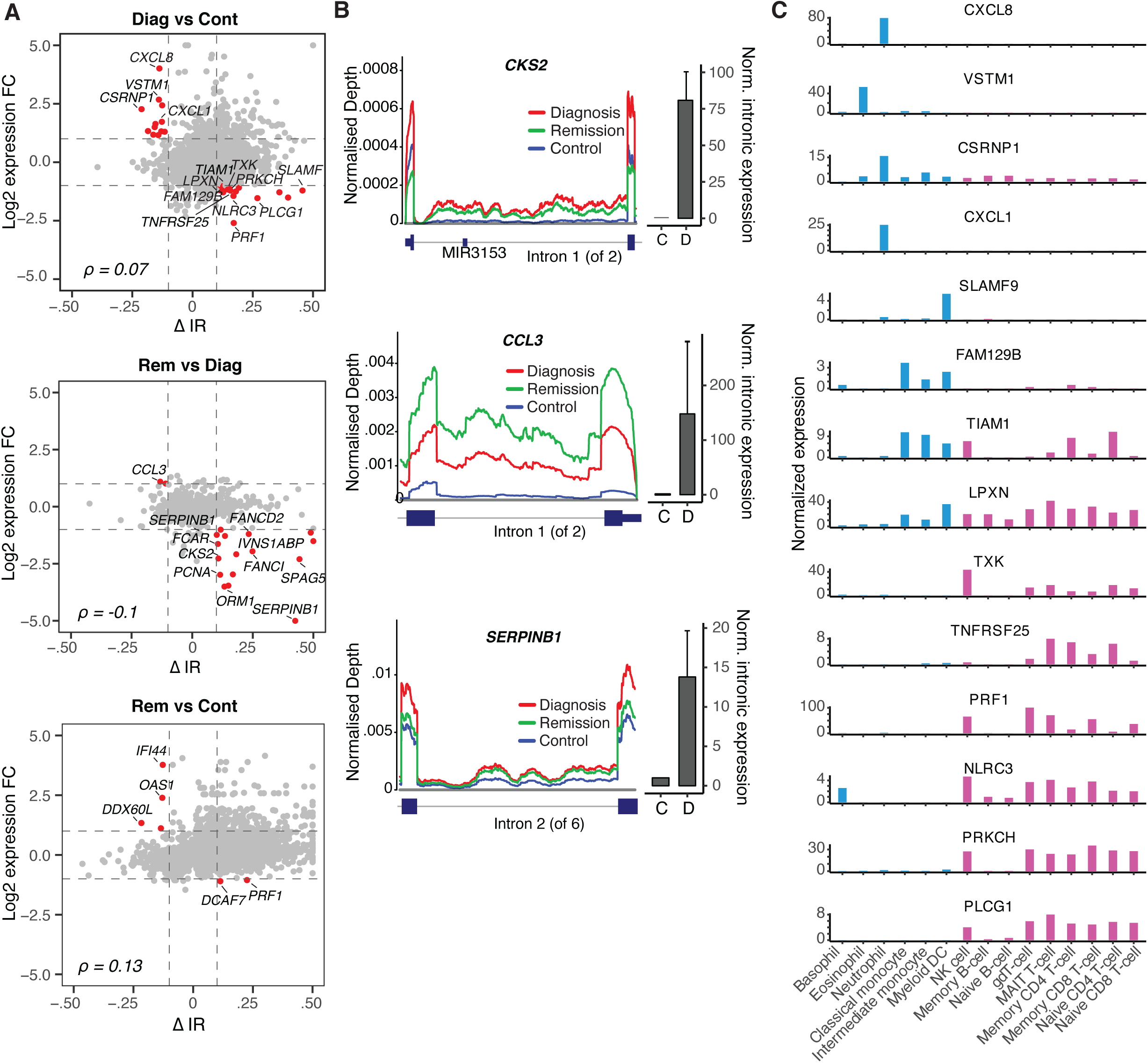
Differentially retained introns inhibit the expression of cell cycle regulators following major molecular remission. **(A)** Scatterplots illustrating the relationship between differentially retained introns (ΔIR) and differential gene expression (log2 expression fold change). Introns (|ΔIR| ≥ 0.1) with inverse relationship to host gene expression (|log(FC)| ≥ 1) are highlighted in red. **(B)** Coverage plots of selected differential IR events (left) and qRT-PCR validation of differential IR between diagnosis and control (right). qRT-PCR validation was performed in triplicates on independent diagnosis samples and results were normalised to *B2M* expression. D - diagnosis; C - control. **(C)** Blood cell type expression of genes with inverse IR/gene expression relationships (diagnosis v control; blue - myeloid, magenta – lymphoid cells). The data was retrieved from the Human Protein Atlas (v19.3; http://www.proteinatlas.org)^49^.

However, many of the genes, whose expression does anti-correlate with the retention of their introns, are involved in cell cycle regulation (e.g. *CCL3, CKS2*, and *SPAG5*), and DNA replication/repair (e.g. *FANCI*/*FANCD2*, and *PCNA*) (Figure 4A).

We selected three differentially retained introns of cell cycle regulator genes for experimental validation (*CKS2* intron 1, *CCL3* intron 1, and *SERPINB1* intron 2). Based on the RNA sequencing data analysis, all three introns have an increased expression and higher IR ratio at diagnosis compared to healthy controls (Figure 4A). We confirmed these observations via qRT-PCR (Figure 4B). The IR ratios of *CKS2* intron 1, and *SERPINB1* intron 2 increase further at remission resulting in a reduction of host gene expression, while the IR ratio of *CCL3* intron 1 is reduced at remission and associated with an increase in gene expression (Figure 4A & B).

Interestingly, some instances of inverse relationships between IR and gene expression affect lineage-specific genes (Figure 4C). For example, the chemokine Interleukin-8 (*CXCL8*), which is predominantly expressed in neutrophils, is upregulated at diagnosis. This could be attributed to the increased myeloid progenitor cell abundance (Figure 1B), but also to the reduced IR levels in *CXCL8* mRNA (Figure 4A). In control samples, the IR levels increase from 12.9% to 26.7% in *CXCL8* (intron 2), while gene expression is reduced 4-fold. *CXCL8* promotes CML cell proliferation^29^ and previous reports suggest that *CXCL8* expression is modulated both by *BCR-ABL1* expression (causing increase) and TKI treatment (causing inhibition of *CXCL8*)^30^. In contrast, *PRKCH* is downregulated at CML diagnosis along with increased IR levels (Figure 4A). *PRKCH* is a serine-threonine kinase that regulates hematopoietic stem cell function and is specifically expressed in lymphoid cells. Notably, high *PRKCH* expression has been associated with poor prognosis in acute myeloid leukemia^31^.

These observations suggest that lineage-specific expression of drivers of cell proliferation and hematopoietic cell differentiation can be mediated through IR level changes in CML.

### Epigenetic features modulate intron retention in CML

Characteristics of the retained introns in our CML cohort are in accordance with previously described attributes, such as their relatively short length (Supplementary Figure 13), and higher GC content (Supplementary Figure 14). While these are intrinsic features of IR, we sought potential *trans*-regulators and found that the gene that most strongly correlates (ρ = 0.64) with IR frequencies is *MED21* (Mediator Complex Subunit 21; Figure 5A). *MED21* regulates gene transcription by interacting with RNA polymerase II^32^. RNA polymerase II elongation rates influence splice site recognition and IR^13^. Interestingly, the expression of *ZNF160*, encoding a Zinc Finger Protein and repressor of transcription, strongly anti-correlates (ρ = −0.62) with IR frequencies (Figure 5A).

**Figure 5.**
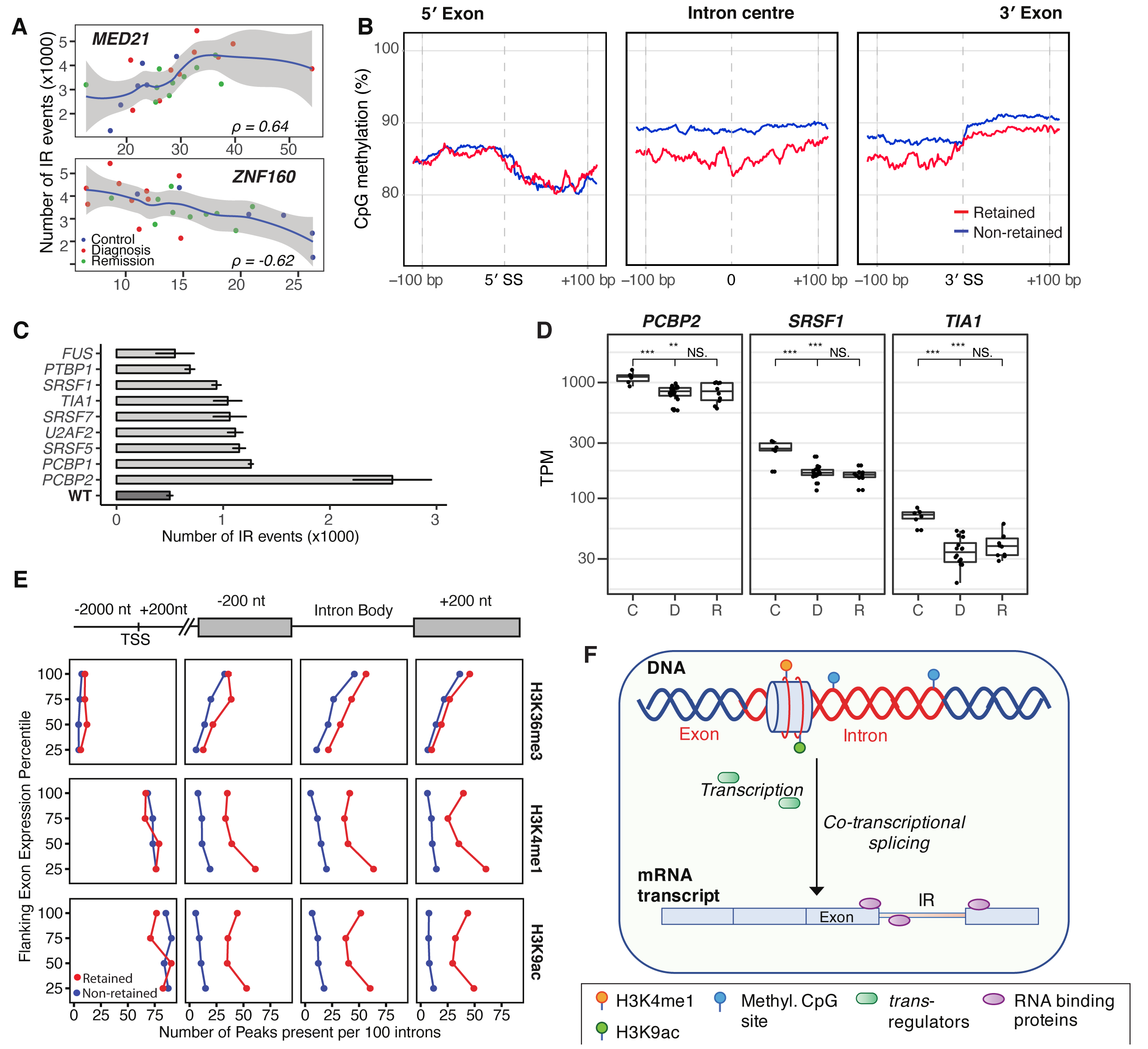
Regulation of IR in CML. **(A)** Two genes that most strongly correlate/anti-correlate with IR frequencies (Spearman correlation). **(B)** CpG methylation around intron splice sites (±200 bp) and in the centre (200 bp) of retained- (red) and non-retained introns (blue) in CML diagnosis samples. General linear hypothesis test; p-value < 2.2e-16 (adj.-p < 1e-10). **(C)** The effect of RNA binding proteins on IR frequency. RNA-seq data of shRNA-mediated knockdown of indicated RNA binding proteins in K562 cells was retrieved from the ENCODE project (encodeproject.org). WT – wild type. **(D)** Expression of RNA binding proteins with enriched binding motifs near frequently retained introns^20^. TPM - transcripts per million mapped reads; C - control; D - diagnosis; R – remission; n.s. – non-significant, * *p* < 0.05, ** *p* < 0.01, *** *p* < 0.001. **(E)** Histone modifications with increased numbers of peaks near splice sites of retained introns (red: retained; blue: non-retained introns). The analysis was repeated for each expression quartile to exclude the possibility that increased histone modifications are associated with gene expression. **(F)** Possible mechanisms of IR regulation in CML discussed in the text.

We have previously shown that DNA methylation regulates IR in myeloid cells^13^. To confirm this mode of regulation in our CML cohort, we compared CpG methylation around the splice sites (±200 bp) and in the centre (200 bp) of retained and non-retained introns. We confirmed decreased DNA methylation levels near the 3□ splice junction and within the body of retained introns (Figure 5B). Similar patterns were observed in remission and control samples suggesting that this mode of IR regulation is also present in lymphoid cells (Supplementary Figure 15).

To further investigate the role of epigenetics in the regulation of IR in CML, we retrieved RNA-seq and ChIP-seq data of the K562 CML cell line from ENCODE. We analysed transcriptomics data of RNA binding protein knockdown experiments and observed significantly increased IR occurrences with the most drastic IR increase (∼5 fold) following Poly(RC) Binding Protein 2 (*PCBP2*) knockdown (Figure 5C). All genes with enriched binding motifs near frequently retained introns^20^ were downregulated at CML diagnosis, except for *SRSF5* (Supplementary Figure 16). Three of these RNA binding proteins, including *PCBP2*, remained downregulated in remission (Figure 5D).

To identify additional potential explanations for the dysregulation of IR in CML, we analysed histone modifications in K562 cells. While higher levels of H3K36 trimethylation have previously been associated with IR^33,34^, we found only marginal differences in H3K36me3 levels in retained versus non-retained introns. Increasing numbers of H3K36me3 peaks around introns were observed in more highly expressed genes (Supplementary Figure 17). However, we found that monomethylation and acetylation of histone H3 lysines 4 and 9, respectively (H3K4me1 and H3K9ac) were significantly enriched near splice sites of retained introns and within their intron bodies in contrast to non-retained introns (Figure 5E). While a significant enrichment of H3K9ac has been found to co-occur at many gene regulatory elements in mouse embryonic stem cells^35^, we here established a strong association of H3K9ac with IR. Elevated levels of H3K4 methylation in association with IR have been previously reported by Zhou et al.^33^.

In summary, our analysis of CML patient transcriptomes revealed aberrant alternative splicing both at diagnosis and remission. Elevated IR ratios in CML can be explained by multiple regulatory mechanisms including the regulation of RNA Pol II elongation, demethylation of CpG sites, and histone modifications (Figure 5F). The TKI treatment itself seems to have no effect on IR (Supplementary Figure 18).

## Discussion

In this study, we analysed epigenomics and transcriptomics data from patients diagnosed with chronic phase Ph+ CML and matched remission samples after effective treatment with TKIs. Thereby, we generated novel insights into the molecular responses to TKI therapy beyond the known reduction in *BCR-ABL1* expression.

We have identified recurring fusion transcripts in patient samples including an uncharacterised fusion between the cell surface marker C-type lectin domain family 12 member A (*CLEC12A*; 12p13.31) and the *MIR223* gene (Xq12). Other non-specific intra-chromosomal fusions include *EEF1DP3-FRY, EIF4E3-FOXP1, KANSL1-ARL17B*, and the mitochondrial fusion *ND6-TE* (Supplementary Figure 2A). An increasing number of mitochondrial mutations and fusions have been described as a consequence of increased reactive oxygen species during ageing^36^, however, the age range (21-70 yrs) of *ND6-TE*-positive patients in our cohort does not support an age-related occurrence of this fusion.

Gene expression profiles of our CML remission samples are more similar to those of healthy donors, likely due to the change in cell composition and the loss of *BCR-ABL1* clones. Consistent with previous studies, we found cell cycle regulators among the most aberrantly expressed genes in the diagnosis samples. For example, cyclins A1 and B2 (*CCNA1, CCNB2*) were among the most differentially expressed genes, with overexpression at diagnosis and low expression at remission (comparable to expression levels observed in healthy controls). While cyclin D2 (*CCND2*) has previously been characterized as a *BCR-ABL*-dependent mediator of cell proliferation in hematopoietic cells^37^, marginal changes in *CCND2* (FC= 0.87, p=.04) expression suggest that other cyclins such as *CCNA1* (FC=12.3, p=5.4e-53) and *CCNB2* (FC=7.2, p= 9.2e-55) were among the more potent regulators of cell proliferation in this cohort of CML patients.

The most surprising results was that, in contrast to gene expression, alternative splicing patterns did not return to normal in CML remission samples. Among other forms of alternative splicing, IR affects transcriptomic complexity and leads to the inclusion of premature termination codons in mature transcripts causing nonsense-mediated decay^38,39,27,39,40^. Normal granulocytic blood cells make use of this conserved mechanism of gene regulation by increasing the number of intron-retaining transcripts of hundreds of genes that are essential to granulocytic differentiation^27,38^. Shen *et al*. have shown that alternative splicing events can be used to construct predictors for patient survival, that outperform gene expression-based predictors in multiple cancers^41^. Splicing-based prognostic markers have already been found for several cancer types^41,42^. Therefore, a splicing-based predictor for TKI cessation success is not just desirable but an achievable prospect, once clinical data of trial TKI cessation together with remission transcriptomics data become available.

Our results were consistent across patients and different specific TKI treatments. It is known that the cell composition of the peripheral blood changes in CML - with myeloid progenitor cell-enrichment at diagnosis and mature lymphoid cell-enrichment at remission. The latter is very similar to the cell composition observed in the peripheral blood of healthy individuals (Supplementary Figure 5). Therefore, epigenomic and transcriptomic differences between diagnosis and remission may be expected. However, remarkable are the persistent alternative splicing perturbations in remission that do not resemble patterns observed in healthy controls.

We found new evidence for epigenetic regulators of alternative splicing, such as reduced DNA methylation, and increased H3K4me1 and H3K9ac marks around retained introns. We have previously shown that DNA methylation can regulate IR in myeloid cells and during terminal granulopoiesis^13,43^. In CML reduced DNA methylation in the intron body and 3□ splice site is associated with IR (Figure 5B). A role for the promoter and enhancer mark H3K4 mono-methylation in gene repression has been identified previously^44^, however, H3K4me1-dependent alternative splicing has not been described before. Both H3K4me1 and H3K9ac are present at the promoters of intron-retaining and non-retaining genes but are predominantly found near splice sites and within bodies of retained introns in contrast to non-retained introns (Figure 5E). While both histone marks seem indicative of active transcription (Figure 5E), by regulating Pol II recruitment and elongation rates^44,45^ around splice sites they could promote IR in CML. IR regulation via H3.3K36me3-specific readers, such as previously shown in HeLa cells^34^, seems less relevant in CML patients.

## Conclusions

In summary, our study has provided new insights into the epigenomic and transcriptomic landscapes of CML patients at diagnosis and remission. We have shown that, despite a changing cell composition at major or complete molecular remission, alternative splicing remains aberrantly regulated with high IR levels affecting multiple cell cycle regulators. Further research is required to test whether sustained RNA processing alterations in CML remission facilitate relapse following TKI cessation. The new knowledge about epigenetic links to alternative splicing in CML could facilitate the development of epigenetic therapeutic adjuvants to increase the likelihood of successful TKI cessation. Modulation of spliceosome function has recently been proposed as a new therapeutic avenue in leukemia^46^, which is an idea reinforced by our results.

## Data Availability

The dataset supporting the conclusions of this article is available in the GEO repository, GSE144119.

https://www.ncbi.nlm.nih.gov/geo/query/acc.cgi?acc=GSE144119

## List of abbreviations

CML: chronic myeloid leukemia
DEU: Differential exon usage
DUEs: differentially used exons
DMRs: Differentially methylated regions
FC: fold change
GO: gene ontology
IR: intron retention
PBMCs: peripheral blood mononuclear cells
Ph+: Philadelphia-positive
TCGA: The Cancer Genome Atlas
TPM: transcripts per million
TKI: tyrosine kinase inhibitor
WGBS: Whole genome bisulfite sequencing

## Declarations

### Ethics approval and consent to participate

All studies were approved by the institutional review boards (HREC protocol No: 131015, 070718c, 081211, and 101010 - Royal Adelaide Hospital) and are in accordance with the Declaration of Helsinki. Patients provided written informed consent.

### Consent for publication

Not applicable.

### Competing interests

S.B. is member of the advisory board of Qiagen, Novartis and Cepheid, received honoraria from Qiagen, Novartis, Bristol-Myers Squibb and Cepheid, and research support from Novartis. D.W. received RS funding and honoraria from BMS and honoraria from Amgen. J.E.J.R. has received honoraria or speakers fees (GSK, Miltenyi, Takeda, Gilead, Pfizer, Spark, Novartis, Celgene, bluebird bio); Director of Pathology (Genea); equity ownership (Genea, Rarecyte); consultant (Rarecyte, Imago); chair, Gene Technology Technical Advisory, OGTR, Australian Government. The remaining authors declare no competing financial interests.

### Funding

This work was supported by the National Health and Medical Research Council (Investigator Grant #1177305 to J.E.J.R., Project #1128175 and #1129901 to J.E.J.R. and J.J.-L.W., #1126306 to J.J.-L.W., APP1027531 and APP1104425 to S.B., and APP1135949 to T.P.H.; the NSW Genomics Collaborative Grant (J.E.J.R. and J.J.- L.W.); Cure the Future (J.E.J.R.); and an anonymous foundation (J.E.J.R.). U.S. and J.J.-L.W. hold Fellowships from the Cancer Institute of New South Wales. U.S. also received support from the Australian Academy of Science in form of an Australia-India Early and Mid-Career Fellowship. Financial support was provided by Tour de Cure (Scott Canner Research Fellowship) to C.G.B. and for research grants to C.G.B. and J.E.J.R. This research was funded by the Cancer Council NSW Project Grants (RG11-12, RG14-09, RG20-12) to J.E.J.R., C.G.B., and U.S. Support was also received from the Ray and Shirl Norman Cancer Research Trust (S.B.).

## Authors’ contributions

J.E.J.R. and J.J.-L.W. designed the research. U.S., J.S.S., P.L.L., and V.P. performed bioinformatic analyses and interpreted the data. D.L.W., V.A.S., S.B., A.G.T., and T.P.H. provided CML patient samples, clinical data and further insight. B.D., G.M., S.N., C.M., and C.G.B. performed and/or supervised experiments. U.S. wrote the manuscript with help from J.J.-L.W., J.S.S., G.M., B.D., and V.P. All authors reviewed and approved the manuscript.

## Acknowledgements

We would like to acknowledge Dr. Andreas Schreiber (University of South Australia) for retrieving the sequencing data from the Branford et al. study^9^.

## References

1. Peng C, Li S. Chronic Myeloid Leukemia (CML) Mouse Model in Translational Research. Methods Mol Biol. 2016;1438:225–243.

2. Talpaz M, Silver RT, Druker BJ, et al. Imatinib induces durable hematologic and cytogenetic responses in patients with accelerated phase chronic myeloid leukemia: results of a phase 2 study. Blood. 2002;99(6):1928–1937.

3. Apperley JF. Part I: mechanisms of resistance to imatinib in chronic myeloid leukaemia. Lancet Oncol. 2007;8(11):1018–1029.

4. Perrotti D, Jamieson C, Goldman J, Skorski T. Chronic myeloid leukemia: mechanisms of blastic transformation. J Clin Invest. 2010;120(7):2254–2264.

5. Yong AS, Melo JV. The impact of gene profiling in chronic myeloid leukaemia. Best Pract Res Clin Haematol. 2009;22(2):181–190.

6. Ma W, Giles F, Zhang X, et al. Three novel alternative splicing mutations in BCR-ABL1 detected in CML patients with resistance to kinase inhibitors. Int J Lab Hematol. 2011;33(3):326–331.

7. Radich JP, Dai H, Mao M, et al. Gene expression changes associated with progression and response in chronic myeloid leukemia. Proc Natl Acad Sci U S A. 2006;103(8):2794–2799.

8. Dvinge H, Bradley RK. Widespread intron retention diversifies most cancer transcriptomes. Genome Med. 2015;7(1):45.

9. Branford S, Wang P, Yeung DT, et al. Integrative genomic analysis reveals cancer-associated mutations at diagnosis of CML in patients with high-risk disease. Blood. 2018;132(9):948–961.

10. Heller G, Topakian T, Altenberger C, et al. Next-generation sequencing identifies major DNA methylation changes during progression of Ph+ chronic myeloid leukemia. Leukemia. 2016;30(9):1861–1868.

11. Wong ACH, Rasko JEJ, Wong JJ. We skip to work: alternative splicing in normal and malignant myelopoiesis. Leukemia. 2018;32(5):1081–1093.

12. Tilgner H, Nikolaou C, Althammer S, et al. Nucleosome positioning as a determinant of exon recognition. Nat Struct Mol Biol. 2009;16(9):996–1001.

13. Wong JJ, Gao D, Nguyen TV, et al. Intron retention is regulated by altered MeCP2-mediated splicing factor recruitment. Nat Commun. 2017;8:15134.

14. Dobin A, Davis CA, Schlesinger F, et al. STAR: ultrafast universal RNA-seq aligner. Bioinformatics. 2013;29(1):15–21.

15. Wang L, Wang S, Li W. RSeQC: quality control of RNA-seq experiments. Bioinformatics. 2012;28(16):2184–2185.

16. Ewels P, Magnusson M, Lundin S, Kaller M. MultiQC: summarize analysis results for multiple tools and samples in a single report. Bioinformatics. 2016;32(19):3047–3048.

17. Haas BJ, Dobin A, Stransky N, et al. STAR-Fusion: Fast and Accurate Fusion Transcript Detection from RNA-Seq. bioRxiv. 2017:120295.

18. Patro R, Duggal G, Love MI, Irizarry RA, Kingsford C. Salmon provides fast and bias-aware quantification of transcript expression. Nature methods. 2017;14(4):417.

19. Shen S, Park JW, Lu ZX, et al. rMATS: robust and flexible detection of differential alternative splicing from replicate RNA-Seq data. Proc Natl Acad Sci U S A. 2014;111(51):E5593–5601.

20. Middleton R, Gao D, Thomas A, et al. IRFinder: assessing the impact of intron retention on mammalian gene expression. Genome Biol. 2017;18(1):51.

21. Anders S, Reyes A, Huber W. Detecting differential usage of exons from RNA-seq data. Genome Res. 2012;22(10):2008–2017.

22. Clark SJ, Statham A, Stirzaker C, Molloy PL, Frommer M. DNA methylation: bisulphite modification and analysis. Nature protocols. 2006;1(5):2353.

23. Nair SS, Luu PL, Qu W, et al. Guidelines for whole genome bisulphite sequencing of intact and FFPET DNA on the Illumina HiSeq × Ten. Epigenetics Chromatin. 2018;11(1):24.

24. Song Q, Decato B, Hong EE, et al. A reference methylome database and analysis pipeline to facilitate integrative and comparative epigenomics. PLoS One. 2013;8(12):e81148.

25. Love MI, Huber W, Anders S. Moderated estimation of fold change and dispersion for RNA-seq data with DESeq2. Genome Biol. 2014;15(12):550.

26. Huang da W, Sherman BT, Lempicki RA. Systematic and integrative analysis of large gene lists using DAVID bioinformatics resources. Nat Protoc. 2009;4(1):44–57.

27. Wong JJ, Ritchie W, Ebner OA, et al. Orchestrated intron retention regulates normal granulocyte differentiation. Cell. 2013;154(3):583–595.

28. Edwards CR, Ritchie W, Wong JJ, et al. A dynamic intron retention program in the mammalian megakaryocyte and erythrocyte lineages. Blood. 2016;127(17):e24–e34.

29. Corrado C, Raimondo S, Saieva L, Flugy AM, De Leo G, Alessandro R. Exosome-mediated crosstalk between chronic myelogenous leukemia cells and human bone marrow stromal cells triggers an interleukin 8-dependent survival of leukemia cells. Cancer Lett. 2014;348(1-2):71–76.

30. Hantschel O, Gstoettenbauer A, Colinge J, et al. The chemokine interleukin-8 and the surface activation protein CD69 are markers for Bcr-Abl activity in chronic myeloid leukemia. Mol Oncol. 2008;2(3):272–281.

31. Porter SN, Magee JA. PRKCH regulates hematopoietic stem cell function and predicts poor prognosis in acute myeloid leukemia. Exp Hematol. 2017;53:43–47.

32. Plaschka C, Lariviere L, Wenzeck L, et al. Architecture of the RNA polymerase II–Mediator core initiation complex. Nature. 2015;518(7539):376.

33. Zhou Y, Lu Y, Tian W. Epigenetic features are significantly associated with alternative splicing. BMC Genomics. 2012;13:123.

34. Guo R, Zheng L, Park JW, et al. BS69/ZMYND11 reads and connects histone H3.3 lysine 36 trimethylation-decorated chromatin to regulated pre-mRNA processing. Mol Cell. 2014;56(2):298–310.

35. Karmodiya K, Krebs AR, Oulad-Abdelghani M, Kimura H, Tora L. H3K9 and H3K14 acetylation co-occur at many gene regulatory elements, while H3K14ac marks a subset of inactive inducible promoters in mouse embryonic stem cells. BMC Genomics. 2012;13:424.

36. Westermann B. Mitochondrial fusion and fission in cell life and death. Nat Rev Mol Cell Biol. 2010;11(12):872–884.

37. Jena N, Deng M, Sicinska E, Sicinski P, Daley GQ. Critical role for cyclin D2 in BCR/ABL-induced proliferation of hematopoietic cells. Cancer Res. 2002;62(2):535–541.

38. Schmitz U, Pinello N, Jia F, et al. Intron retention enhances gene regulatory complexity in vertebrates. Genome Biol. 2017;18(1):216.

39. Wong JJ, Au AY, Ritchie W, Rasko JE. Intron retention in mRNA: No longer nonsense: Known and putative roles of intron retention in normal and disease biology. Bioessays. 2016;38(1):41–49.

40. Kurosaki T, Maquat LE. Nonsense-mediated mRNA decay in humans at a glance. J Cell Sci. 2016;129(3):461–467.

41. Shen S, Wang Y, Wang C, Wu YN, Xing Y. SURVIV for survival analysis of mRNA isoform variation. Nat Commun. 2016;7:11548.

42. Li Y, Sun N, Lu Z, et al. Prognostic alternative mRNA splicing signature in non-small cell lung cancer. Cancer Lett. 2017;393:40–51.

43. Gao D, Pinello N, Nguyen TV, et al. DNA methylation/hydroxymethylation regulate gene expression and alternative splicing during terminal granulopoiesis. Epigenomics. 2019;11(1):95–109.

44. Cheng J, Blum R, Bowman C, et al. A role for H3K4 monomethylation in gene repression and partitioning of chromatin readers. Mol Cell. 2014;53(6):979–992.

45. Gates LA, Shi J, Rohira AD, et al. Acetylation on histone H3 lysine 9 mediates a switch from transcription initiation to elongation. J Biol Chem. 2017;292(35):14456–14472.

46. Lee SC, Dvinge H, Kim E, et al. Modulation of splicing catalysis for therapeutic targeting of leukemia with mutations in genes encoding spliceosomal proteins. Nat Med. 2016;22(6):672–678.

47. Aran D, Hu Z, Butte AJ. xCell: digitally portraying the tissue cellular heterogeneity landscape. Genome Biol. 2017;18(1):220.

48. Krzywinski M, Schein J, Birol I, et al. Circos: an information aesthetic for comparative genomics. Genome Res. 2009;19(9):1639–1645.

49. Uhlen M, Oksvold P, Fagerberg L, et al. Towards a knowledge-based Human Protein Atlas. Nat Biotechnol. 2010;28(12):1248–1250.

